# Analysis of temporal trends in potential COVID-19 cases reported through NHS Pathways England

**DOI:** 10.1101/2020.05.16.20103820

**Authors:** Quentin J. Leclerc, Emily S. Nightingale, Sam Abbott, CMMID COVID-19 Working Group, Thibaut Jombart

**Affiliations:** Department of Infectious Disease Epidemiology, London School of Hygiene and Tropical Medicine, Keppel Street, London, UK; Department of Global Health and Development, London School of Hygiene and Tropical Medicine, Keppel Street, London, UK; UK Public Health Rapid Support Team, London, UK; MRC Centre for Global Infectious Disease Analysis, Department of Infectious Disease Epidemiology, School of Public Health, Imperial College, London, UK

## Abstract

The NHS Pathways triage system collates data on enquiries to 111 and 999 services in England. Since the 18th of March 2020, these data have been made publically available for potential COVID-19 symptoms self-reported by members of the public. Trends in such reports over time are likely to reflect behaviour of the ongoing epidemic within the wider community, potentially capturing valuable information across a broader severity profile of cases than hospital admission data. We present a fully reproducible analysis of temporal trends in NHS Pathways reports until 14th May 2020, nationally and regionally, and demonstrate that rates of growth/decline and effective reproduction number estimated from these data may be useful in monitoring transmission. This is a particularly pressing issue as lockdown restrictions begin to be lifted and evidence of disease resurgence must be constantly reassessed. We further assess the correlation between NHS Pathways reports and a publicly available NHS dataset of COVID-19-associated deaths in England, finding that enquiries to 111/999 were strongly associated with daily deaths reported 16 days later. Our results highlight the potential of NHS Pathways as the basis of an early warning system. However, this dataset relies on self-reported symptoms, which are at risk of being severely biased. Further detailed work is therefore necessary to investigate potential behavioural issues which might otherwise explain our conclusions.

## Introduction

The coronavirus 2019 disease (COVID-19) is caused by SARS-CoV-2, a novel coronavirus originally identified in Wuhan (China) in December 2019 [1]. As of the beginning of May 2020, more than 3 million confirmed COVID-19 cases and 200,000 deaths have been reported worldwide [1]. To mitigate epidemic spread and reduce the risk of healthcare systems being overrun, countries have implemented different measures to slow down transmission, such as travel restrictions and various degrees of social distancing [2].

More than 120,000 confirmed COVID-19 cases and 25,000 deaths have been reported in England as of the beginning of May 2020 [3]. However, laboratory confirmation has largely been restricted to hospitalised cases [3]. The majority of milder cases, which likely represent a large fraction of the total are unaccounted for [4]. We should therefore explore alternative, community-driven surveillance data to help characterise the progression of the COVID-19 epidemic in the country.

NHS Pathways is a triage system for public calls and online reports for medical care [5]. This system is currently being used throughout England to assist individuals reporting potential COVID-19 symptoms. Since the 18th of March 2020, data on daily phone calls and completed online assessments which have received a potential COVID-19 final disposition are [openly available](https://digital.nhs.uk/data-and-information/publications/statistical/mi-potential-covid-19-symptoms-reported-through-nhs-pathways-and-111-online/latest). These assessments are either completed via calls to 111 and 999 (which are respectively for non-urgent, and urgent medical problems), or through 111-online self-completed reports. The fraction of assessments corresponding to actual COVID-19 cases is unknown, but in the absence of wide-scale testing, the NHS Pathways dataset may be one of the best available proxies for COVID-19 incidence in the community. While prone to self-reporting biases, it is likely to better reflect milder cases and be less biased by different severity profiles than hospital admission data, which by definition reflect the most acute cases.

Here, we analyse NHS Pathways data until 14th May 2020 to assess the temporal dynamics of COVID-19 in England. Specifically, we investigated potential changes in the growth rate of the epidemic over time, and compared the observed patterns across NHS regions. We derived time-varying estimates of the growth rates, halving time and effective reproduction numbers for the different regions. We also assessed the potential correlation between NHS Pathways data with COVID-19 daily deaths in England, to gain an initial understanding of its possible value within an early detection system.

## Methods

### Data extraction

We extracted the NHS Pathways data up to 14th May 2020 through the NHS Digital website [5], where they are updated daily, every weekday. This dataset contains daily numbers of calls to 111 and 999, as well as 111-online reports that were classified as potential COVID-19 cases by the triage algorithm. This algorithm follows a symptom-based approach, where the respondents are asked a series of questions to self-report their symptoms until an end point is reached, or the call is handed to a clinician [6]. Some individuals might report symptoms multiple times through the system, therefore the number of reports are not directly equivalent to the underlying number of cases. The number of reports are stratified by Clinical Commissioning Group (CCG), gender and age group (0–18, 19–69 and 70–120 years old) of the patients. We mapped the CCGs to their corresponding NHS regions using publicly available CCG data [7], and used this geographic resolution for our analysis. All dates indicated refer to the date of reporting.

A reporting change in the NHS Pathways data occured over the period between the 9th and 23rd of April, during which the number of 111-online reports for individuals aged between 0 and 18 years old was not available. However, due to the relatively small proportion of total reports for that age group, this likely does not affect our results (see Figures S3-S5 in *Supplementary Material* for further details). We also note that towards the end of March 2020, patients with potential COVID-19 symptoms were directed away from HP services, which may have increased 111 reporting.

### Temporal analysis

Total numbers of reports (including all three data sources: 111 calls, 999 calls, and 111-online reports) were modelled using quasi-Poisson generalised linear models (GLM) with log links, to account for exponential trends as well as over-dispersion of the data [8]. Predictors included time (in days since the first data point (18th March 2020) with interaction terms for varying slopes and intercepts between NHS regions, and day of the week (weekend, monday, or rest of the week) to account for potential differences in reporting over the weekend and at the start of the week. Growth rates (*r*) for each NHS region and their 95% confidence intervals were directly deduced from the corresponding coefficients. All models were fitted using maximum-likelihood.

To assess potential changes of the growth rate over time, analyses were performed over rolling windows of 14 days from the earliest available date (18th March 2020) to the latest available one (14th May 2020; see Figure S1 in *Supplementary Material* for a sensitivity analysis). Growth rates and associated confidence intervals were calculated for each time window. Whenever the upper bound of *r* was negative, corresponding halving times were calculated as *log(0.5)/r*. Growth rates were converted to effective reproduction numbers *R_e_* using the approach described in Wallinga and Lipsitch [9] and implemented in the *epitrix* package [10], with a serial interval modelled as a gamma distribution with mean 4.7 days and standard deviation 2.9 days [11] (see Figure S2 in *Supplementary Material* for further details on the choice of distribution).

### Correlation with reported deaths

To test the validity of the NHS Pathways dataset as an early detection system, we compared daily total counts of reports (including all three data sources: 111 calls, 999 calls, and 111-online reports) with publicly available NHS data on COVID-19 daily deaths [12]. This dataset includes daily counts of COVID-19 deaths in hospitals in England NHS regions. All dates refer to the date of death. However, the data are subject to bias from reporting delays, with more recent counts excluding a proportion of deaths which have not yet been reported. To account for this, we excluded data from the last 3 weeks (i.e. from the 23rd April 2020) from this analysis.

We calculated Pearson’s correlation between the daily time series of deaths and NHS Pathways reports, lagging the reports from zero to thirty days. Approximate 95% confidence intervals for each correlation estimate were calculated by bootstrapping with 1,000 replicates. From this we identified an optimal lag at which the reports correlate most strongly with subsequent deaths. We then further evaluated the potential of NHS Pathways reports lagged at this value as a predictor, assuming a quasi-Poisson distribution for daily deaths.

All analyses were performed using the R software [13], and the code is publicly available from https://github.com/qleclerc/nhs_pathways_report and distributed under the MIT license.

## Results

Overall reports of potential COVID-19 cases through NHS Pathways have been clearly decreasing in all NHS regions since 18th March 2020 and until approximately 13th April 2020, after which the trend seems to plateau (Figure 1). Weekly spikes were consistently observed for all NHS regions, with increased reports on Mondays likely reflecting less reporting over weekends. The NHS COVID-19 daily deaths dataset shows daily deaths increased until approximately 10th April 2020, and have been decreasing since.

**Figure 1.**
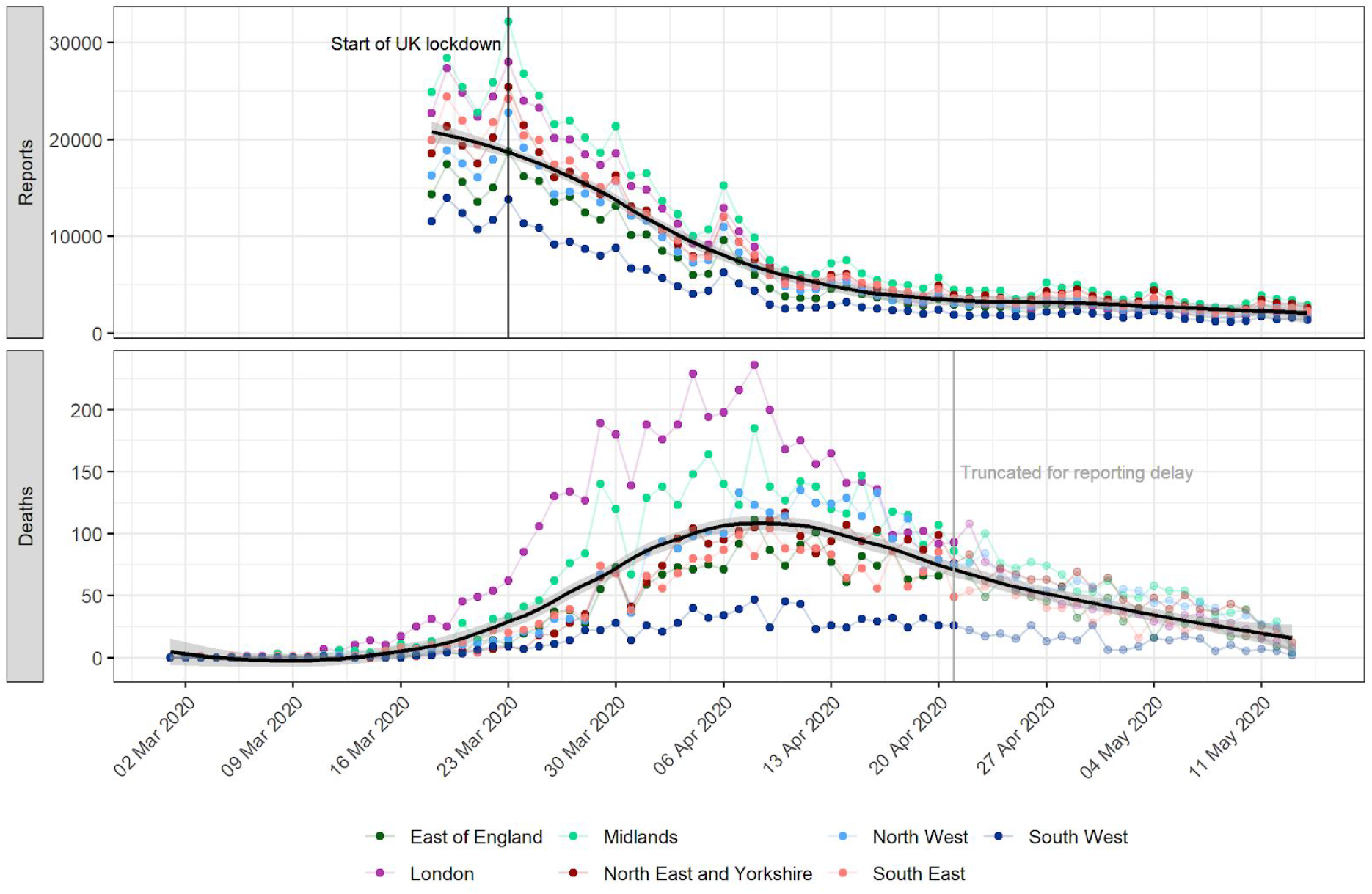
Daily potential COVID-19 cases reported through NHS Pathways and reported COVID-19-related deaths, by NHS region. Pathways data include calls to 111 and 999, as well as 111-online reports. Dates correspond to the date of case report and death report, respectively, with x-axis labels corresponding to Mondays. The solid black line and grey ribbon correspond to a lowess smoother and its 95% confidence interval. The start of the lockdown in England (23rd March 2020) and date at which death data were truncated to avoid bias from reporting delay (21st April 2020) are highlighted by vertical lines.

As epidemics are expected to decrease exponentially, a plateau in case incidence would be expected even if the decline rate (i.e., negative growth rate *r*) remained constant over the time period considered. However, this could also reflect a genuine change in *r* over time. Analyses over sliding time windows show that daily decline rates have likely been changing substantially during this period. Results show a marked decrease in *r* and the corresponding effective reproduction numbers (*R_e_* until the 6th April 2020, after which these numbers remained low in all NHS regions for a period of about two weeks (Figure 2). The lowest *r*_*e*_) until the 6th April 2020, after which these numbers estimated was for Saturday 18 April 2020 in the London region at –0.08 (95% CI: –0.10 –−0.06), corresponding to a halving time of 8.45 days (95% CI: 6.77 – 11.20) and an *R_e_* of 0.66 (95% CI: 0.59 – 0.73). Similar trends were observed across all NHS regions, with the exception of London which showed lower *r* (and *R_e_*) after the 13th of April.

**Figure 2.**
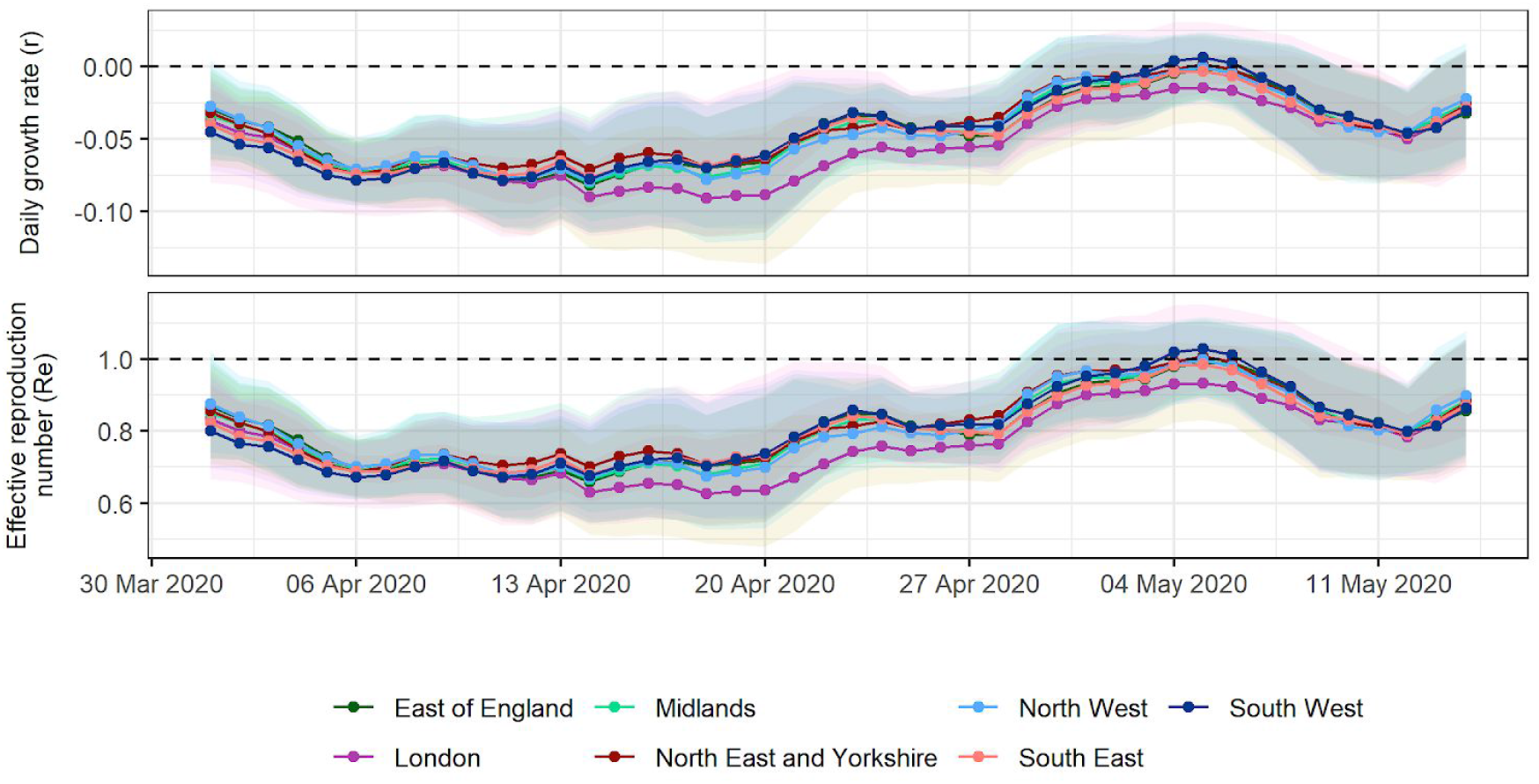
Estimates of daily growth rates (r) and effective reproduction numbers (R_e_) for potential COVID-19 cases reported through NHS Pathways. Dotted lines indicate the central estimate, and ribbons their 95% confidence intervals. Estimates are indicated at the end of the time window used for estimation, so that values of r and R_e_ provided on a given day correspond to the 2 weeks leading up to that day.

Confidence intervals suggest *r* values remained lower than 0 (and *R_e_* lower than 1) in all regions until 15th April, consistent with a decrease in COVID-19 related reports. After this, values of *r* and *R * seem to have gradually increased, to a point where there was no longer any strong evidence of a decrease in COVID-19 reports in any NHS region as of 2nd May 2020, as the 95% CIs of all growth rates include 0. The most recent estimate of *r* averaged over all NHS regions is –0.03 (95% CI: –0.06 – 0.01), corresponding to an *R_e_* of 0.88 (95% CI: 0.73 – 1.04).

The strongest correlation between NHS Pathways reports and deaths was obtained with a lag of 16 days (Pearson’s correlation = 0.94; 95% CI: 0.78 – 0.98). Figure 3 illustrates the observed trend in correlation across all tested lags. Estimates become increasingly unstable for lags above 25 days as the number of points within the overlapping time window becomes small (*n* = 5 at 30 days lag). There is however a clear, initially upward trend and subsequent plateau between 16 and 19 days, after which the strength of correlation appears to decrease. Further analysis suggests that this pattern is also observed when looking at NHS regions separately, albeit with some variation in the optimal lag (see Figure S6 in *Supplementary Material* for further details).

**Figure 3.**
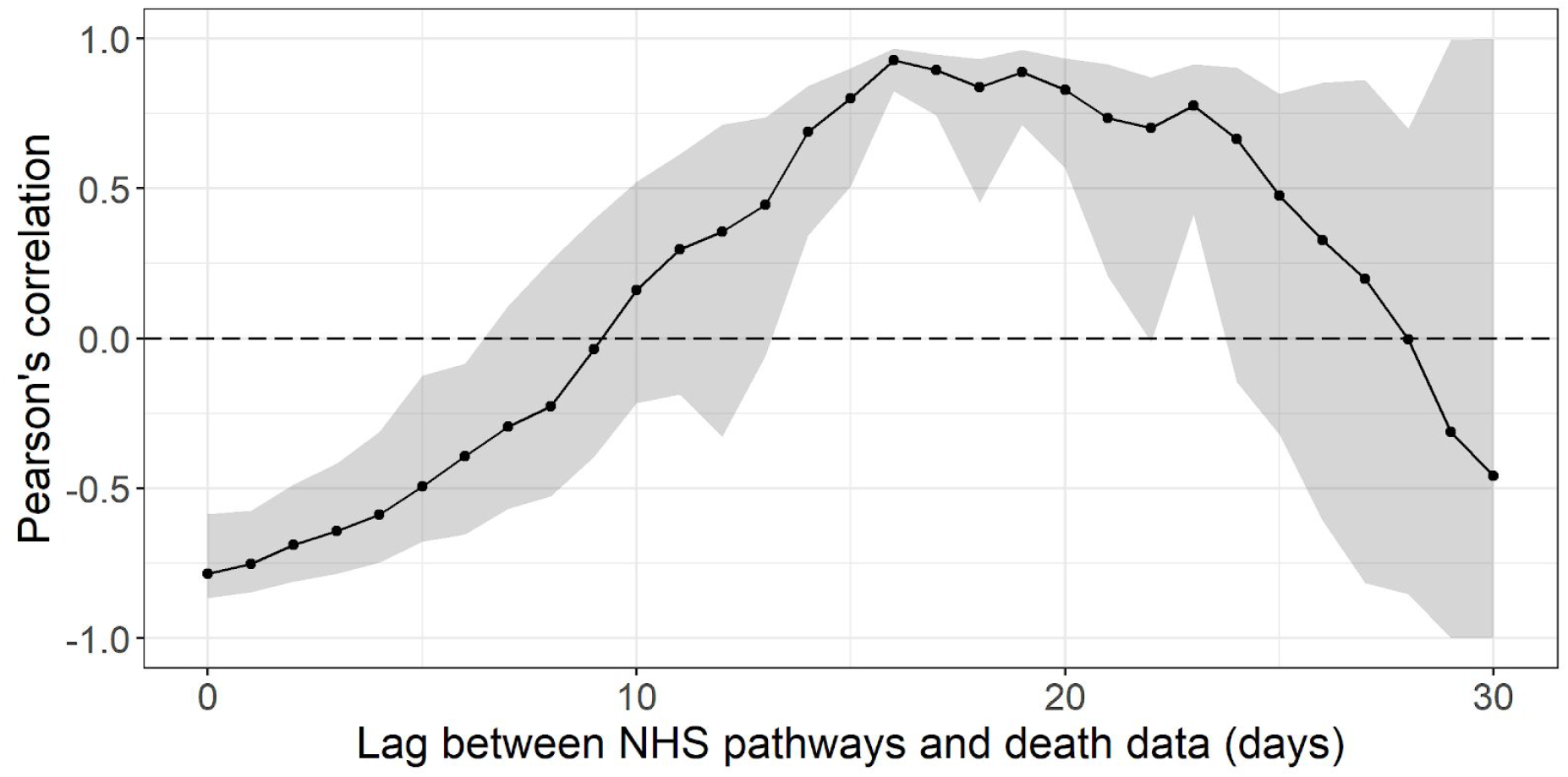
Pearson’s correlation between deaths and potential COVID-19 cases reported through NHS Pathways, lagged between 0 and 30 days. 95% confidence intervals are calculated by bootstrapping with 1,000 replicates.

Fitting a quasi-Poisson GLM, we found that over 85.8% of the deviance in daily reported deaths could potentially be explained by NHS Pathways reports 16 days prior, with an average of 1.91 (bootstrap 95% CI: 1.70 – 2.07) additional deaths for every 1,000 potential COVID-19 cases reported in NHS Pathways 16 days before (intercept = 397, 95% CI: 357 – 442; % increase per 1000 notifications = 0.48, 95% CI: 0.39 – 0.57; Figure 4).

**Figure 4.**
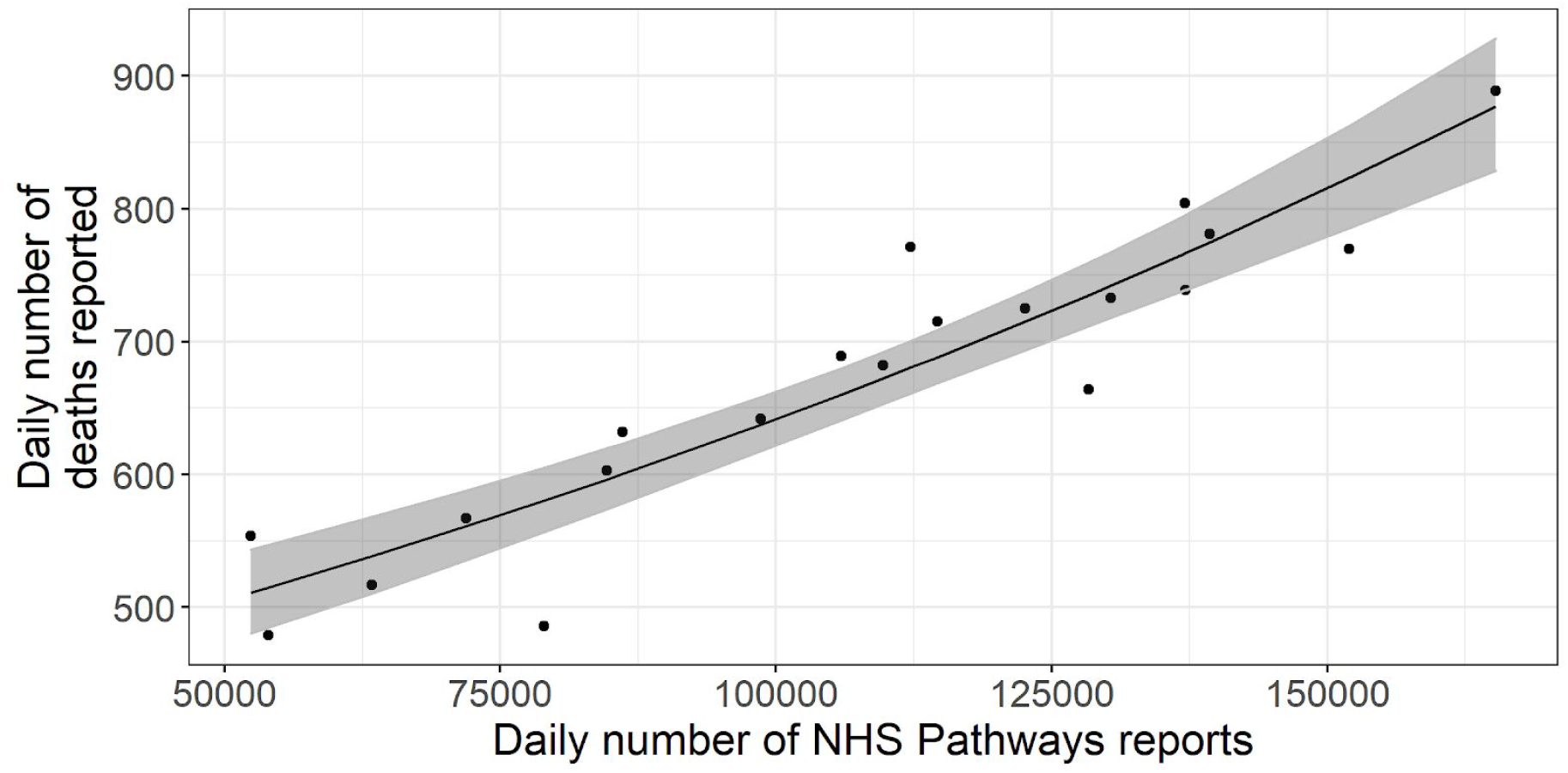
Daily total COVID-19 deaths reported in England between 3rd and 22nd April 2020, against the number of potential COVID-19 cases reported through NHS Pathways with a lag of 16 days (between 18th March and 6th April 2020). The black line and grey ribbon correspond to predictions from the regression model and associated 95% confidence intervals. The coefficient of determination indicates that 85.8% of the deviance in reported deaths is linearly explained by NHS Pathways reports.

## Discussion

We analysed publicly available NHS Pathways data to assess the temporal dynamics of the COVID-19 epidemic in England until 14th May 2020. Trends in NHS Pathways reports are similar across all England NHS regions, based on casual observation. Our results suggest after a sharp initial decrease up until early April 2020, transmissibility may have slowly increased until early May. After this, it becomes unclear if cases are still declining as confidence intervals of *r* and *R_e_* include the threshold values for growth / decline (respectively, 0 and 1) in all NHS regions. Trends in COVID-19 associated deaths in England showed more variation between NHS regions than the Pathways reports, with one potential explanation being differing fatality risk amongst cases due to age and comorbidities. As a national total, the number of reported deaths was found to be strongly correlated with the number of reported 16 days prior. However, if these data are to serve as an early warning for potential disease resurgence, further investigation will be required to ascertain whether this correlation holds beyond this period of decline in both trends which we have analysed here.

A number of caveats may affect our results. Firstly, the data we considered are at best a proxy for the true incidence of COVID-19 in the country as they rely on self-reported symptoms interpreted by an algorithm, and were not confirmed by virological tests. This is further exacerbated by the fact that individuals might access NHS Pathways more than once to report their symptoms, which could artificially increase the numbers of potential COVID-19 cases.

As NHS Pathways is based on self-reporting, several biases could affect the data, such as changes in service availability and delays in the uptake of the 111-online reporting system. When estimating growth rates over time and geographic regions, we implicitly assume that self-reporting behaviours have not substantially changed over time, and have been similar across different NHS regions. In reality, self-reporting could be strongly biased by behavioural issues, such as the effect of news coverage which might lead individuals to pay more attention to their symptoms and report them. Inversely, individuals could reduce their perception of the risk of the disease over time as they become used to hearing about it daily, which would decrease their likelihood of noticing and reporting symptoms. Similarly, differences in self-reporting behaviours across various age groups would likely bias the age composition of the potential COVID-19 cases reported here.

While NHS Pathways data may better capture the epidemic in the community than hospitalisation data, these data do not only reflect community cases. In fact, a fraction of cases reported through 999 are likely to be hospitalised, as well as smaller proportions of cases reported through 111 calls and 111-online. Therefore, the data we considered here most likely reflect the epidemic as a whole, rather than just in the community. We also note that all results presented rely on dates of calls or online reports, so that estimates of transmissibility are likely lagged by a few days compared to the true, underlying epidemic. Unfortunately, the delays from exposure to notification through NHS Pathways cannot be estimated from current publicly available data.

The analysis of lagged correlation between the NHS Pathways and deaths data is limited by the observed time windows. With a longer window, correlation estimates for higher lags would be more stable and perhaps the optimal lag would be clearer. However, a recent review found that estimates of hospital length of stay from admission to death were mostly below 10 days [14], therefore a potential lag of 16 days between potential COVID-19 cases reported through NHS Pathways and COVID-19 deaths would be coherent with the timelines for patients observed so far. Future work will aim to better exploit the temporal correlation in these data by regressing on a series of lags.

The initial sharp decrease in potential COVID-19 cases could suggest that social distancing measures put in place have had a strong impact on reducing transmission and brought the effective reproduction number under 1 across all NHS regions. However, the more recent estimates of *R_e_* suggest that transmissibility might have increased again since, such that it is no longer clear that case incidence is declining. The main limitation in interpreting this result is that as true incidence reaches low levels in the population, the relative proportion of false positives among potential COVID-19 cases reported through NHS Pathways is likely to increase. In fact, if there were no more cases but an approximately constant number of false positives, the analysis of NHS Pathways data would estimate *R_e_* to be around 1. Nevertheless, the observed changes in estimated transmissibility over time, together with the potentially strong correlation between NHS Pathways reports and COVID-19 deaths time series suggest that future changes in *R_e_* could possibly be reflected in NHS Pathways data. Moreover, we found that, for the time period covered by our analysis, the NHS Pathways data were a good predictor of COVID-19 deaths reported 16 days later, making this data source a good candidate for designing early warning systems in the upcoming weeks as lockdown restrictions are progressively lifted in England. Further work is now needed to test whether this correlation holds outside a declining epidemic, and to investigate potential behavioural issues which might otherwise explain the trends we have highlighted in our analysis.

## Data Availability

All analyses were performed using the R software [13], and both the code and data are publicly available from https://github.com/qleclerc/nhs_pathways_report and distributed under the MIT license.

## Supplementary material

### Sensitivity analysis for rolling window model

A rolling window analysis allows us to better capture trends in the data, by reducing the impact of noise between different days on our estimates. However, the choice of the time window can affect our estimates. Using a 7 days window in our model leads to high variation between estimates (Figure S1A), while a 21 days window prevents us from capturing finer trends in the data (Figure S1B). We therefore chose to report results for a 14 days window as a compromise between these two sizes.

**Figure S1.**
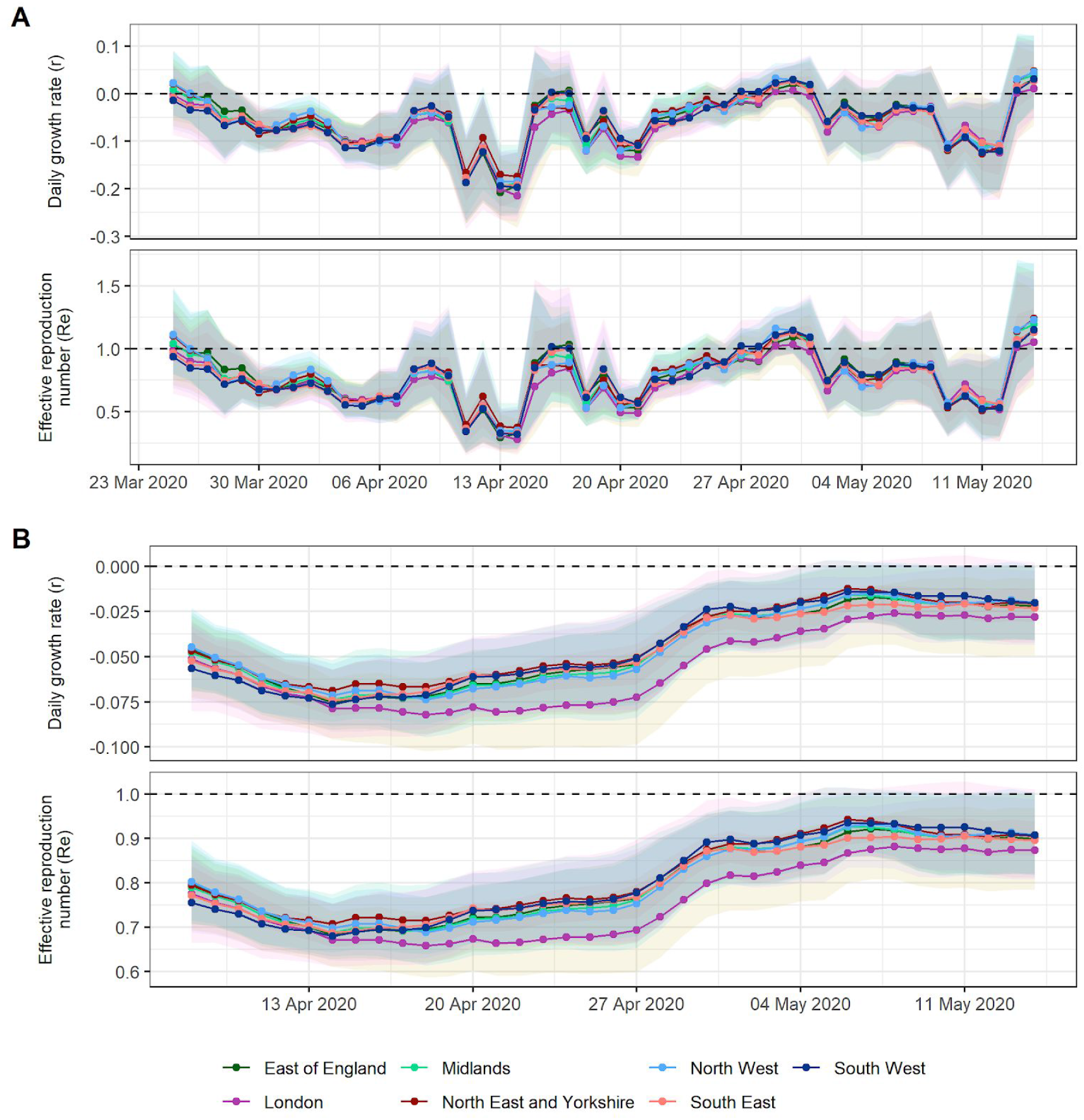
Estimates of daily growth rates (r) and effective reproduction numbers (R_e_ for potential COVID-19 cases (all ages) reported through NHS Pathways. Dotted lines indicate the central estimate, and ribbons their 95% confidence intervals. Estimates are indicated at the end of the time window used for estimation, so that values of r and R_e_ provided on a given day correspond to the chosen number of weeks leading up to that day. The size of the rolling window is A) 1 week; and B) 3 weeks.

### Serial interval distribution

We parameterised our serial interval distribution with a mean of 4.7 days and a standard deviation of 2.9 days. These parameters were obtained from a previous study by Nishiura et al [11], where the authors concluded that a lognormal distribution provided the best fit to their data. However, in our analysis the non-truncated lognormal distribution led to high serial interval values, with a maximum value greater than 200 days (Figure S2B), which heavily influenced our *R_e_* estimates. We therefore chose to use a gamma distribution instead, which generated a more appropriate range of serial interval values for our analysis (Figure S2A).

**Figure S2.**
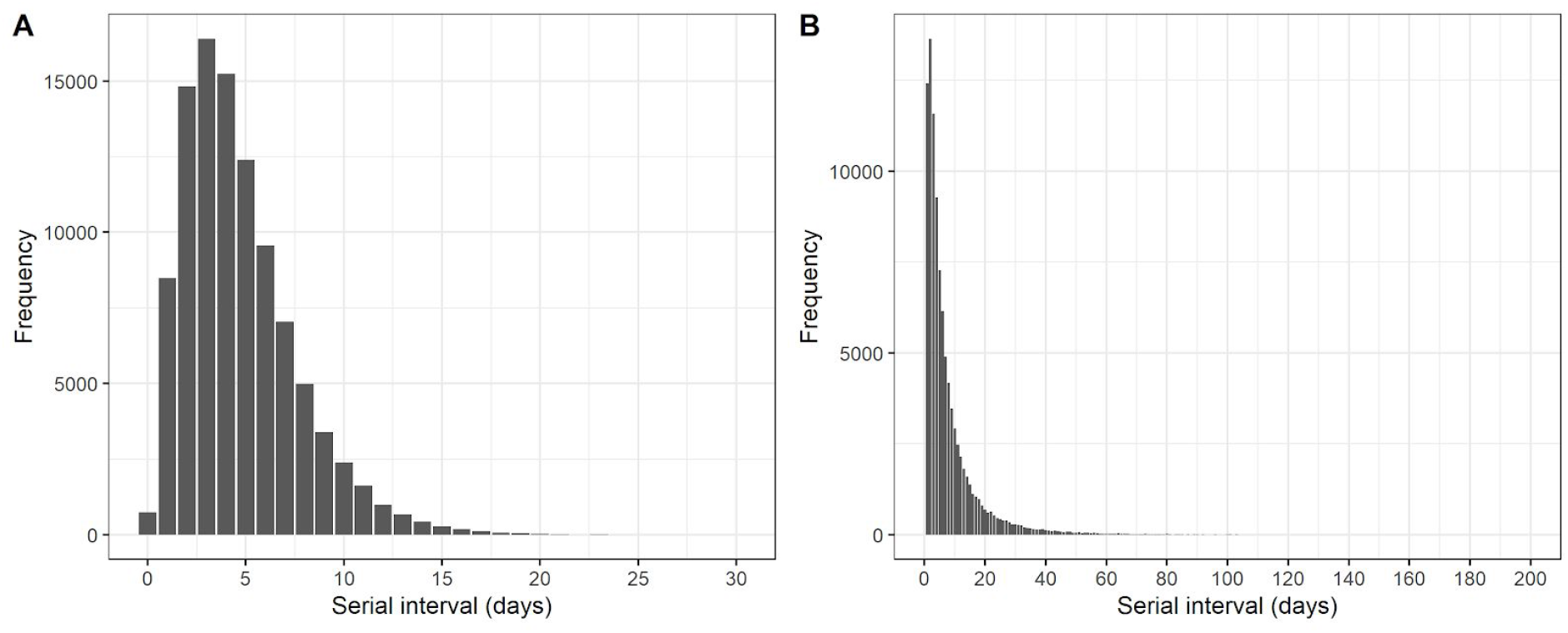
Serial interval distributions considered for our analysis. A) Gamma distribution; B) Lognormal distribution. Both distributions are parameterised with a mean of 4.7 days and a standard deviation of 2.9 days. Serial interval values above 200 are not plotted.

### Age patterns

Reports are classified into 3 age groups in the NHS Pathways dataset: 0–18, 19–69 and 70–120 years old. A majority of all reports (86%) came from individuals between 19 and 69 years old (Figures S3 and S4). Despite some temporal variation, the proportion of cases from this age group exceeded proportions in the general population in all NHS regions (Figure S4). The proportion of reports of potential cases aged 0–18 years old decreased rapidly in all regions from mid-March (about 20%) to early April (about 10%). We note that the proportion of reports for 0–18 years old was lowest between the 9th and 23rd April, which corresponds to the period where reports from this age group were not included in the 111-online subset of the data. However, since this age group represents a minority of total reports, and reports for other age categories were also decreasing at that time (Figure S5), this lack of reporting is likely not responsible for the overall negative growth rates we presented in our results.

**Figure S3.**
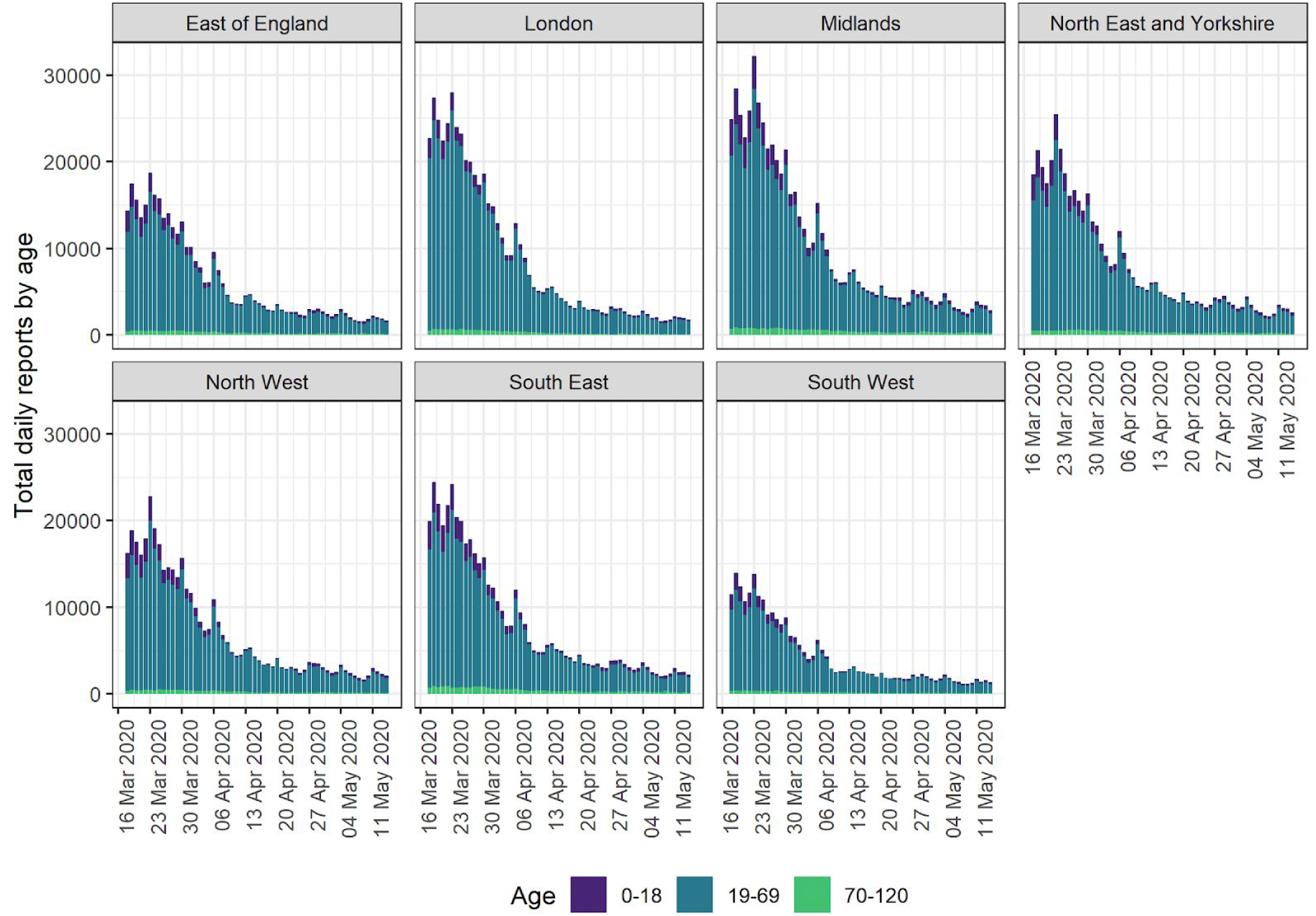
Daily potential COVID-19 cases reported through NHS Pathways, by NHS region and age group. Data include calls to 111 and 999, as well as 111-online reports. Dates correspond to the date of case report, with x-axis labels corresponding to Mondays.

**Figure S4.**
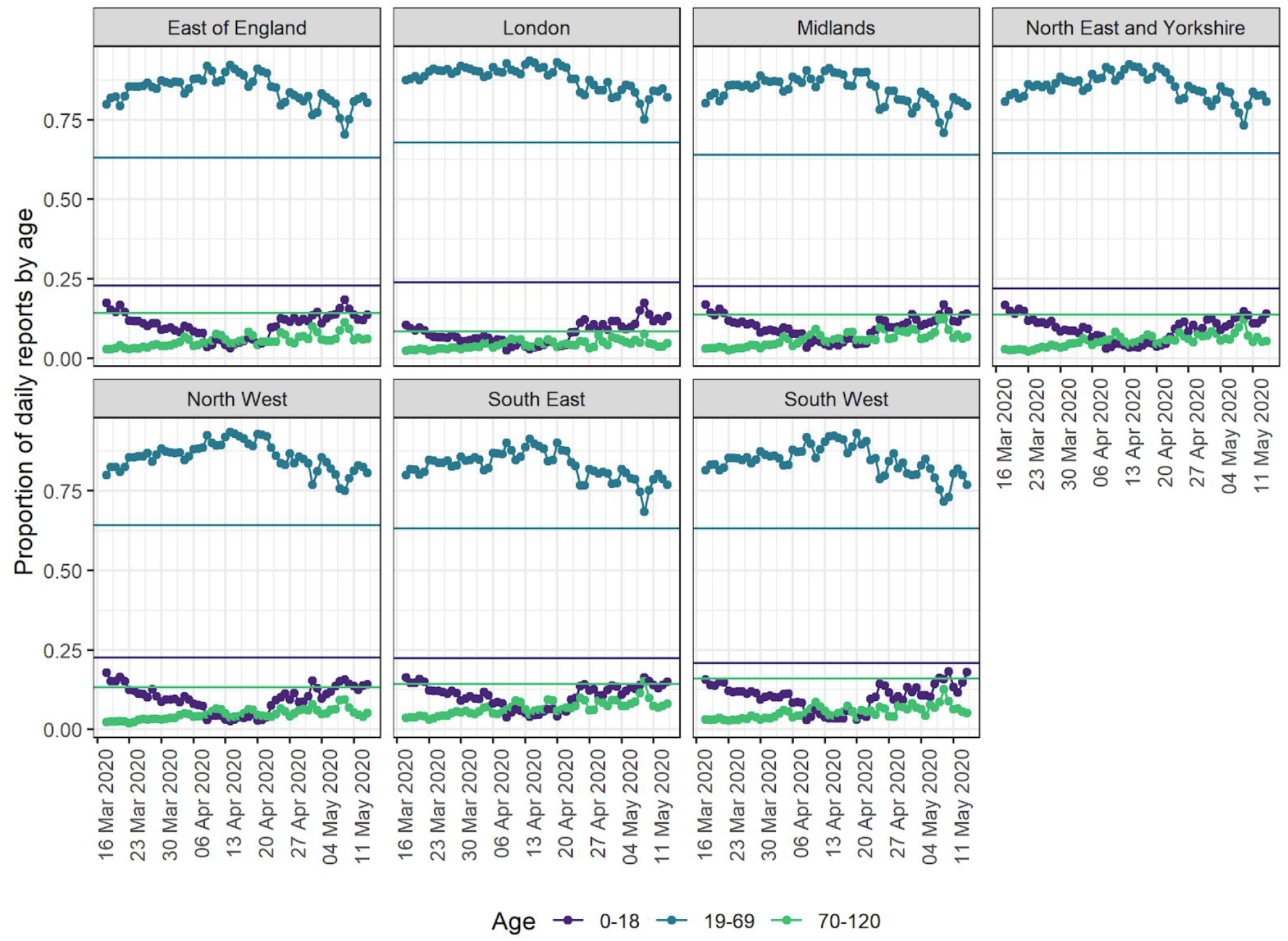
Proportion of potential COVID-19 cases reported through NHS Pathways by age group. Dates correspond to dates of report. Proportions are derived from all potential COVID-19 reports including 111 and 999 calls, and 111-online. Horizontal, solid lines represent the age distribution for the total population of the corresponding NHS region.

**Figure S5.**
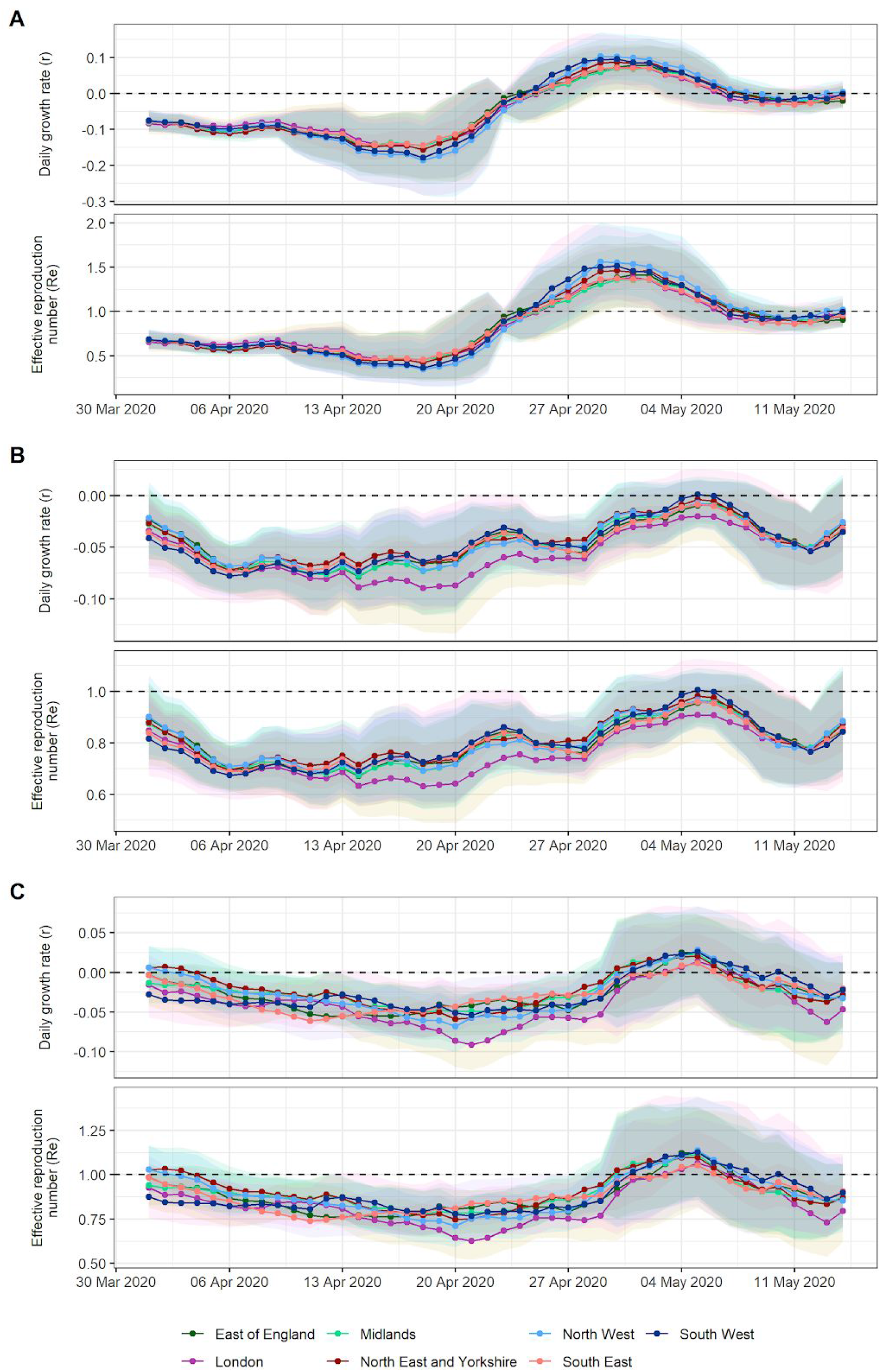
Estimates of daily growth rates (r) and effective reproduction number (R_e_) for potential COVID-19 cases reported through NHS Pathways, stratified by age. A) Cases aged 0 to 18 years old; B) Cases aged 19 to 69 years old; C) Cases aged 70 to 120 years old. Lines and points indicate the central estimate, and ribbons their 95% confidence intervals. Estimates are indicated at the end of the time window used for estimation, so that values of r and R_*e*_ provided on a given day correspond to the 2 weeks leading up to that day.

### Correlation between NHS Pathways reports and deaths by region

We repeated the correlation analysis between NHS Pathways reports and deaths for each region separately. The values suggest that the correlation reported in the main text for England overall could also potentially hold true for trends seen at the regional level, but the corresponding lag time would vary by a few days between regions (Figure S6). At this stage however, we cannot conclude this with certainty, as the correlation values are further from 1 than the one we report in the main text for England overall.

**Figure S6.**
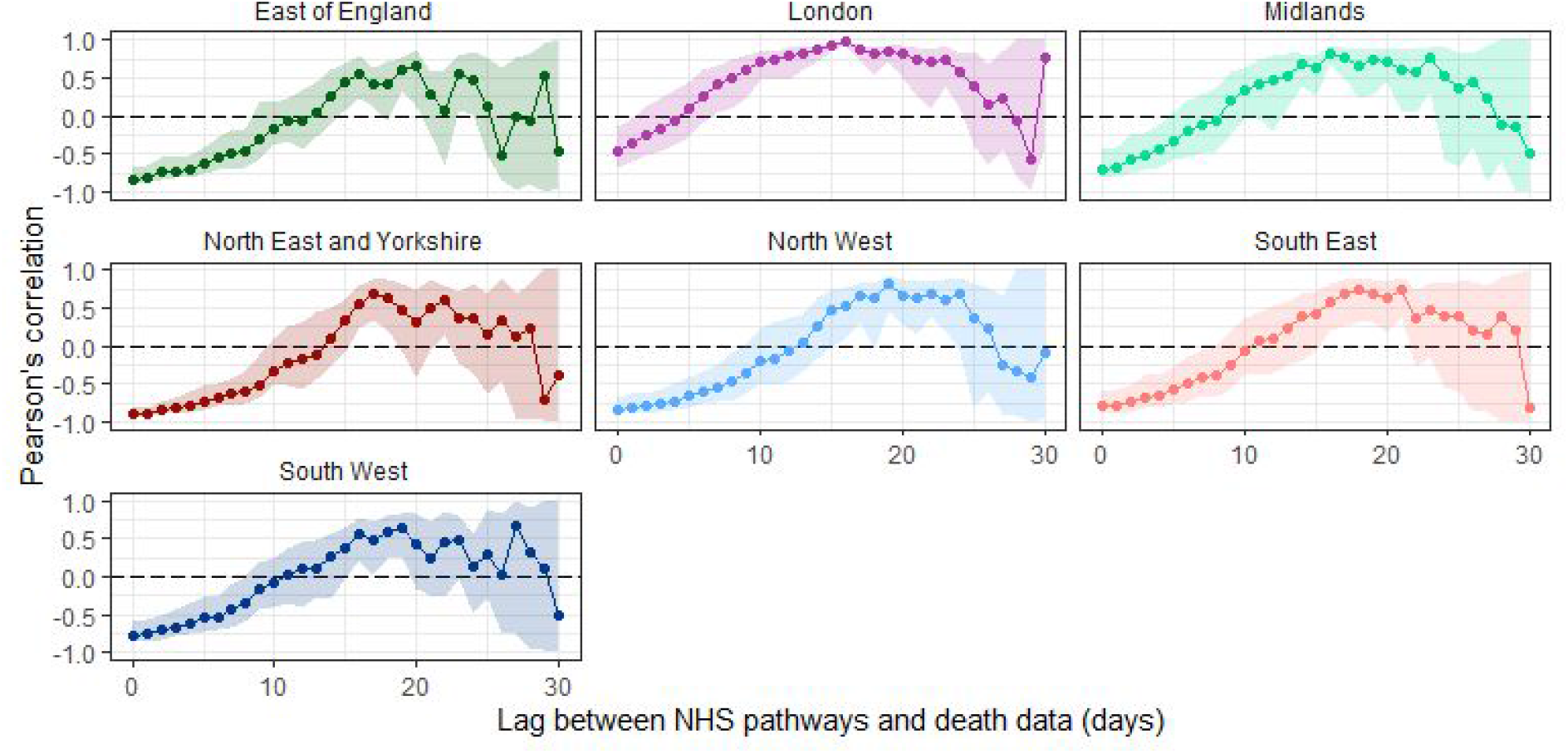
Pearson’s correlation between deaths and potential COVID-19 cases reported through NHS Pathways, lagged between 0 and 30 days and separated by NHS regions. 95% confidence intervals are calculated by bootstrapping with 1,000 replicates.

## References

1. World Health Organization. Coronavirus disease (COVID-19) Situation Report-105 [Internet]. Available from: https://www.who.int/docs/default-source/coronaviruse/situation-reports/20200504-covid-19-sitrep-105.pdf?sfvrsn=4cdda8af{\_}2

2. A third of the global population is on coronavirus lockdown — here’s our constantly updated list of countries locking down and opening up [Internet]. [cited 2020 May 5]. Available from: https://www.businessinsider.fr/us/countries-on-lockdown-coronavirus-italy-2020-3

3. Coronavirus (COVID-19) cases in the UK [Internet]. [cited 2020 May 5]. Available from: https://coronavirus.data.gov.uk/

4. Mizumoto K, Kagaya K, Zarebski A, Chowell G. Estimating the asymptomatic proportion of coronavirus disease 2019 (COVID-19) cases on board the Diamond Princess cruise ship, Yokohama, Japan, 2020. Eurosurveillance [Internet]. 2020 Mar;25(10):2000180. Available from: https://www.eurosurveillance.org/content/10.2807/1560-7917.ES.2020.25.10.2000180

5. Potential Coronavirus (COVID-19) symptoms reported through NHS Pathways and 111 online-NHS Digital [Internet]. [cited 2020 May 5]. Available from: https://digital.nhs.uk/data-and-information/publications/statistical/mi-potential-covid-19-symptoms-reported-through-nhs-pathways-and-111-online/latest

6. NHS Pathways-NHS Digital. NHS Digital [Internet]. [cited 2020 May 13]. Available from: https://digital.nhs.uk/services/nhs-pathways

7. Clinical Commissioning Group to NHS England (Region, Local Office) and NHS England (Region) (April 2019) Lookup in England | Open Geography portal [Internet]. [cited 2020 May 5]. Available from: http://geoportal.statistics.gov.uk/datasets/clinical-commissioning-group-to-nhs-england-region-local-office-and-nhs-england-region-april-2019-lookup-in-england?page=6

8. McCullagh P, Nelder J A. Generalized Linear Models, Second Edition. 1989. CRC Press.

9. Wallinga J, Lipsitch M. How generation intervals shape the relationship between growth rates and reproductive numbers. Proceedings of the Royal Society B: Biological Sciences. 2007 Feb;274(1609):599–604.

10. Jombart T, Cori A. Epitrix: Small helpers and tricks for epidemics analysis [Internet]. 2019. Available from: https://CRAN.R-proiect.org/package=epitrix

11. Nishiura H, Linton NM, Akhmetzhanov AR. Serial interval of novel coronavirus (COVID-19) infections. International Journal of Infectious Diseases. 2020 Apr;93:284–6.

12. NHS UK. COVID-19 Daily Deaths [Internet]. [cited 2020 May 13]. Available from: https://www.england.nhs.uk/statistics/statistical-work-areas/covid-19-daily-deaths/

13. R Core Team. R: A language and environment for statistical computing [Internet]. Vienna, Austria: R Foundation for Statistical Computing; 2020. Available from: https://www.R-proiect.org/

14. Rees EM, Nightingale ES, Jafari Y, Waterlow N, Clifford S, Jombert T, Procter SR, Knight GM, CMMID Working Group. COVID-19 length of hospital stay: a systematic review and data synthesis. medRxiv. 2020 Jan.

